# Genomic Surveillance Reveals Clusters of *Plasmodium falciparum* Antimalarial Resistance Markers in Eswatini, a Low-Transmission Setting

**DOI:** 10.1101/2025.07.30.25332463

**Authors:** Nomcebo Nhlengethwa, Andrés Aranda-Díaz, Sibonakaliso Vilakati, Alfred Hubbard, Isobel Routledge, Sonja B. Lauterbach, Takalani I. Makhanthisa, Faith De Amaral, Mukosha Chisenga, Brighton Mangena, Chadwick Sikaala, John Chimumbwa, Quinton Dlamini, Jaishree Raman, Jennifer L. Smith, Sabelo Vusi Dlamini

## Abstract

Elimination of *Plasmodium falciparum* malaria in Eswatini remains elusive due to ongoing importation and sustained local transmission. The current status of antimalarial and diagnostic resistance in the country remains unknown. Genomic surveillance can complement routine surveillance by characterizing the prevalence and distribution of resistance markers and revealing granular patterns of transmission.

Between March and December 2023, dried blood spots, demographic data, and travel history were collected from individuals testing positive for malaria by rapid diagnostic test (RDT), and symptomatic RDT-negative individuals across Eswatini. Multiplexed amplicon deep sequencing was used to genotype 12 genes associated with antimalarial resistance, detect *hrp2*/*3* deletions, estimate genetic relatedness, and detect non-*falciparum* species.

Data from 437 samples revealed significant clustering of parasites carrying resistance markers. A validated marker of artemisinin partial resistance, *kelch13* P553L, was identified in a cluster of 4 infections (0.9% of all samples), likely linked to importation. The *dhps*/*dhfr* sextuple mutant haplotype, associated with high-level sulfadoxine-pyrimethamine resistance, was found in 7.8% of the samples, also within clusters. The *mdr1* N86 genotype associated with reduced lumefantrine susceptibility was nearly fixed. Over 60% of the infections were polyclonal, with higher complexity observed in imported cases. No *hrp2*/*3* deletions were detected; most false-negative RDTs were attributed to subpatent parasitemia. Non-*falciparum* co-infections were very rare (<1%).

These findings shed light on the dynamics of resistance emergence in low-transmission settings. Integrating routine genomic surveillance into national malaria programs is essential to detect, track, and respond to resistance threats in countries nearing elimination.

## Introduction

Malaria elimination in Eswatini, a southern African country targeting elimination by 2025, remains elusive, in part due to persistent local transmission and frequent importation of cases, which fuel periodic upsurges.^1^ Although national malaria prevalence remains very low, malaria still affects hundreds of individuals each year.^1^ Between 2021 and 2022, Eswatini reported an average of 470 malaria cases per year, including 79 severe cases and 3 deaths annually (Eswatini National Malaria Programme [NMP], personal communication, 2025). To curb transmission, Eswatini has implemented a combination of interventions targeting both the vector and the parasite, including indoor residual spraying (IRS) and prompt treatment with artemisinin-based combination therapies (ACTs). The effectiveness of the interventions targeting the parasite depends on timely case detection and efficacious treatment. However, this effectiveness is increasingly threatened by the emergence and spread of *Plasmodium falciparum* strains resistant to antimalarial drugs and diagnostics across Africa.^2,3^

Artemisinin partial resistance (ART-R), characterized by delayed parasite clearance following ACT treatment, once confined to the Greater Mekong Subregion, has independently emerged in countries in East and the Horn of Africa.^4–6^ While resistance to ACT partner drugs has not been confirmed in Africa as it has in Southeast Asia, increasing reports of ACT treatment failures and reduced partner drug susceptibility in Africa are concerning.^7–9^ Additionally, *P. falciparum* strains undetectable by the widely used falciparum-specific histidine rich protein 2 (HRP2)-based rapid diagnostic tests (RDTs) are now prevalent in the Horn of Africa.^10,11^ These developments underscore the need to closely monitor antimalarial drug and diagnostic resistance to ensure continued progress towards elimination.

Antimalarial drug resistance is mediated by mutations in the parasite genome that reduce drug sensitivity or impair the parasite’s clearance rate, potentially leading to treatment failure. ART-R is associated with single nucleotide polymorphisms (SNPs) in the *kelch13* gene, particularly amino acid substitutions in the propeller domain of the Kelch13 protein.^12^ Over the last decade, SNPs in the *kelch13* gene with clinical and/or laboratory evidence of ART-R have been increasingly reported in Ethiopia, Eritrea, Tanzania, Uganda, and Rwanda.^10,13–17^ Mutations in other genes, including the chloroquine resistance transporter (*crt*) and multidrug resistance 1 (*mdr1*), are associated with reduced sensitivity to the ACT partner drugs, lumefantrine, piperaquine, and amodiaquine.^18,19^ Copy number variations in the *plasmepsin 2*/*3 and mdr1* genes, though less common in Africa, have been associated with piperaquine and mefloquine resistance, respectively.^20,21^ Resistance to sulfadoxine and pyrimethamine results from the accumulation of mutations in the dihydropteroate synthase (*dhps*) and dihydrofolate reductase (*dhfr*) genes, which are widely prevalent across Africa and show distinct geographic patterns.^22–24^ Chloroquine resistance, primarily mediated by the *crt* K76T genotype and modulated by additional *crt* and *mdr1* mutations, has declined following chloroquine withdrawal as a first-line *falciparum* malaria treatment in Africa.^25–30^

Accurate diagnosis is critical for effective case management and detecting index cases for reactive interventions. Deletions in the genes coding for histidine-rich proteins 2 and 3 (*hrp2* and *hrp3*) compromise the performance of HRP2-based RDTs, and are prevalent in Ethiopia and Eritrea.^10,11,31^ False-negative RDT results may also arise from parasite densities below the limit of detection, common in low transmission settings, or from high parasite loads that cause a prozone-like effect.^32,33^

Malaria molecular surveillance can serve as an early warning system by detecting hotspots of antimalarial drug and/or diagnostic resistance, guiding the implementation of therapeutic efficacy surveys, and informing treatment policy in areas with declining drug efficacy.^34^ Genomic technologies, including targeted amplicon deep sequencing, provide a cost-efficient method of generating molecular data from samples collected through passive or active surveillance, especially when integrated into routine surveillance platforms.^35^ The only malaria molecular surveillance study in Eswatini, conducted in 2010, found mutation prevalence consistent with widespread historical use of chloroquine and sulfadoxine-pyrimethamine (SP).^36^ Since adopting artemether-lumefantrine as first-line treatment for uncomplicated malaria in 2010, no follow-up investigations or therapeutic efficacy studies have been conducted.^37^

In addition to drug and diagnostic resistance monitoring, parasite genetics can be exploited to estimate measures of within-host and population-level genetic diversity that can provide insights into transmission intensity and dynamics.^38–41^ When linked to case data, parasite genetic relatedness can be estimated to reveal granular patterns of local transmission, identify clusters of closely related infections, and help reconstruct transmission chains.^42–45^

To provide an updated overview of antimalarial drug and diagnostic resistance marker prevalence in Eswatini, we conducted malaria parasite genomic surveillance as part of the Genomics for Malaria in the Elimination 8 (GenE8) initiative - a regional effort aimed at strengthening malaria genomic surveillance in five Southern African countries (Angola, Eswatini, Namibia, South Africa, and Zambia). Here, we report the prevalence of antimalarial resistance markers in Eswatini and characterize clusters of infections carrying resistance markers using parasite genomics, demographic characteristics, and host risk factors.

## Materials and Methods

### Study site

This study included all four administrative regions of Eswatini, a landlocked country bordered by Mozambique to the east and South Africa to the north, west, and south (**Figure 1A**). Eswatini has a population of approximately 1.2 million, with around 30% residing in malaria-receptive areas -defined by the abundant presence of malaria vectors, *Anopheles* mosquitoes, and ecological and climatic factors that favor malaria transmission - located mainly in the lowveld areas of the eastern Lubombo and Hhohho regions.^1,46,47^ Malaria transmission is seasonal, typically occurring between September and May, with most cases reported between January and May.^1^ The annual parasite index for the 2021-2022 season was approximately 0.7 cases/1000 population (NMP, personal communication, 2025). The predominant malaria parasite species is *P*. *falciparum*, with non-*falciparum* infections rarely detected.^48,49^ The primary mosquito vector is *Anopheles arabiensis*.^1^

**Figure 1.**
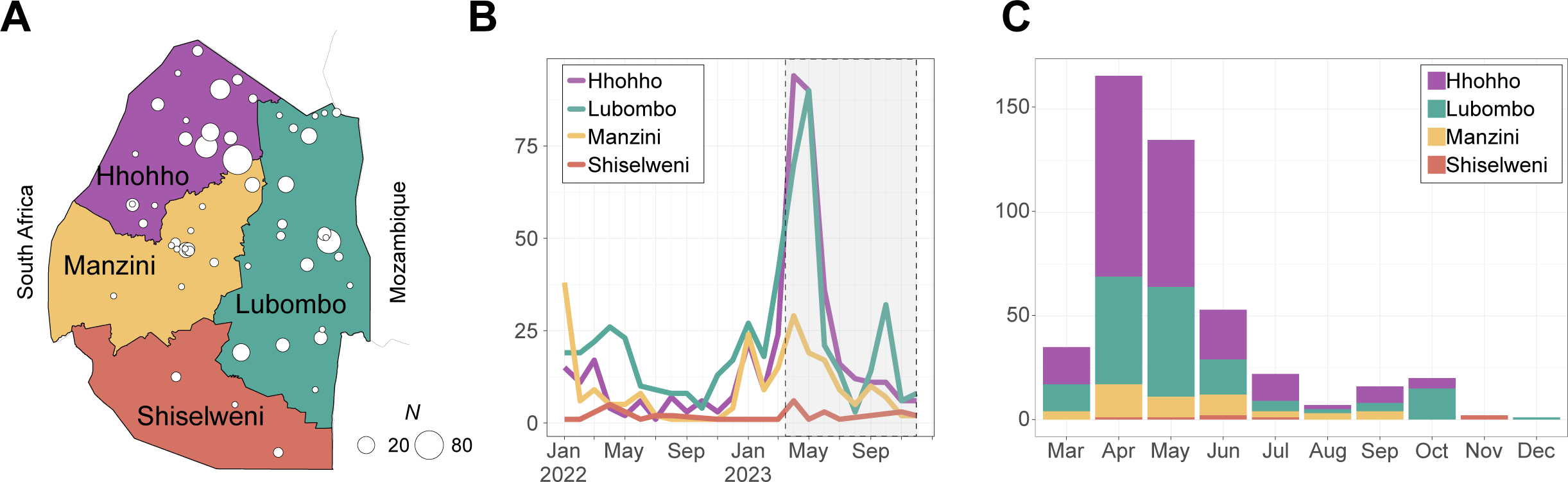
Study Site. **A**. Map of Eswatini and health facilities with sequenced samples. **B**. Total number of reported malaria cases in Eswatini in 2022 and 2023. The study period for this report is highlighted in a dashed box. **C**. Number of sequenced samples stratified by month.

Of the 365 health facilities in Eswatini’s healthcare system, 265 offer malaria diagnosis and treatment services. Diagnosis is primarily based on Pf/Pan RDT (Parascreen, Zephyr Biomedicals, or Meriscreen, Meril Diagnostics) that detects *P*. *falciparum* HRP2 and pan-*Plasmodium* lactate dehydrogenase (pLDH) antigens. Microscopy is reserved for confirmation at facilities with laboratory capacity.^48^ Uncomplicated malaria cases are treated with artemether-lumefantrine combined with a single low dose of primaquine, while severe cases are treated with intravenous or intramuscular artesunate. Mefloquine- or atovaquone-proguanil-based chemoprophylaxis is provided to travelers going to endemic countries at no cost.

Proactive and reactive case detection (RACD) is conducted in receptive areas. Proactive case detection during outbreaks is targeted to high-risk locations such as farms, border posts, and identified transmission hotspots, and is performed based on available programmatic resources and competing priorities. In 2022, 845 individuals were screened by proactive case detection, yielding 1 RDT-positive case (NMP, personal communication 2025). RACD is routinely carried out in all households located within a 500-meter radius of the household of an index case. Eswatini does not currently implement mass drug administration campaigns. In 2022, 59 index cases triggered testing of 1,958 individuals, yielding 28 RDT-positive cases (NMP, personal communication 2025).

Beyond case management, the country’s malaria elimination strategies include annual targeted IRS in homesteads located within receptive areas. Reactive spraying is conducted in the household of every index case located in receptive areas, irrespective of prior household spraying. Distribution of long-lasting insecticidal nets is limited to high-risk groups not protected by IRS (i.e., persons engaged in agricultural activities in the forests).

### Study Design and Field Procedures

This cross-sectional study was conducted as part of the Regional GenE8 (ReGenE8) project and was integrated into Eswatini’s routine malaria surveillance system. All participants aged over 2 years who tested positive for malaria by Pf/Pan RDT or microscopy through passive, reactive, or proactive case detection between March and December 2023 were targeted for inclusion and deep sequencing (**Figure 1**). Based on historical data (590 cases in 2019), up to 900 malaria-positive samples were anticipated for sequencing. An additional 200 individuals testing negative for malaria by RDT but displaying symptoms consistent with malaria were targeted to assess *hrp2*/*3* deletions. Children aged less than 2 years old and individuals who had received treatment or chemoprophylaxis in the preceding 14 days were excluded from the study.

Written informed consent was obtained from all participants prior to enrollment, as described in the Ethical Considerations section. As part of routine case investigation activities, enrolled participants were interviewed using an electronic questionnaire to collect demographic data, clinical symptoms, travel history, and occupation. Collected data was uploaded to the NMP’s Malaria Surveillance Database System, which serves as a central repository for all malaria-related intervention data. Capillary blood samples were collected via finger-prick on filter paper strips (Cytiva Whatmann 3MM Chr), labeled with unique barcodes linking survey and laboratory data. Dried blood spots (DBS) were air-dried, stored in hard paper cards within plastic bags with desiccant at room temperature, and transported to the NMP laboratory in Manzini within a week of collection. In instances where DBS collection was not possible, the RDT cassette was labeled and stored in the same conditions. All samples were stored at −20 degrees in the NMP laboratory before being shipped to the regional sequencing hub at the National Institute for Communicable Diseases in South Africa.

### Sample Processing and Genomic Assays

Parasite densities of genomic DNA extracted from a 6 mm DBS disc or the RDT nitrocellulose strip using the Chelex-Tween 20 method, were estimated using the varATS qPCR assay.^50,51^ All samples with >100 parasites/μL of blood and a random selection of samples with parasitemia between 10 and 100 parasites/μL of blood were selected for sequencing.

Amplicon sequencing libraries were prepared using the MAD4HatTeR targeted amplicon sequencing panel primer pools D1.1, R1.2, and R2.1.^52^ Libraries were sequenced on a NextSeq 2000 instrument using 150-cycle paired-end reads. A custom bioinformatics pipeline was used to infer alleles at 241 targets.^52^ This report includes SNPs and microhaplotypes (SNPs within the same amplicon) from the following genes: *kelch13*, *crt*, *mdr1*, multidrug resistance 2 (*mdr2)*, *dhps*, *dhfr*, *coronin*, apicoplast ribosomal protein S10 (*arps10*), putative phosphoinositide-binding protein (*pib7*), ferrodoxin (*fd*), exonuclease (*exo)*, and PF3D7_1322700. Lactate dehydrogenase (*ldh*) gene target sequences in *P*. *falciparum*, *P*. *vivax*, *P*. *malariae*, *P*. *ovale* and *P*. *knowlesi* were used to identify the presence of *Plasmodium* species in the samples. Targets in and around the *hrp2* and *3* genes were used to identify gene deletions. Microhaplotype sequences from 165 highly heterozygous targets were used to infer complexity of infection (COI) and genetic relatedness. Variations to the original protocol and bioinformatics pipeline are described in Eloff, Aranda-Díaz et al, 2025.^53^

### Data Cleaning, Analysis, and Visualization

Data cleaning, analysis, and visualization were conducted in R (University of Auckland, New Zealand, version 4.3.1). Samples with <10,000 reads, targets with <50 reads, and SNPs with within-sample allele frequency (WSAF) <0.01 or with ≤10 reads were excluded from the analysis. SNPs observed in only one sample with WSAF <1, and SNPs observed in multiple samples but exclusively in a single sequencing run, were also excluded. Library preparation batches with high contamination in negative or positive controls were excluded.

Non-*falciparum* targets were excluded from sequencing depth summaries. Otherwise, all resulting data were analyzed, including samples with partial SNP coverage, leading to variable total sample numbers for each assessed SNP. Samples were considered successfully sequenced if a genotype was obtained for all 59 SNPs evaluated in the *kelch13* gene. Multilocus haplotypes for polymorphisms in 5 independent targets in *dhps* and *dhfr* genes (*dhps* 436, 437, 540, 581, and 613; and *dhfr* 51, 59, 108, and 164) were inferred for each sample by combining all individual microhaplotypes. Haplotypes were inferred for samples with either unmixed genotypes at all targets or with a single target showing a mixed genotype, in which case the sample was assumed to carry the two haplotypes accounting for the observed mixture. Samples with mixed genotypes in more than two amplicons were considered indeterminate.

To assess data patterns and identify potential issues with sampling and library preparation, sample similarity was estimated using the root mean square error of WSAF for highly diverse loci (pool D1.1 targets), followed by Density-Based Spatial Clustering of Applications with Noise (DBSCAN) with ε=0.2. Samples with clonality ≥2 and nearly identical WSAF across all loci, suggesting contamination or sample duplication, were excluded from the analysis.

### Statistical Analysis

Descriptive statistics for gender, age group (<5, 5-14, 15-24, 25-39, 40-59, >59 years), occupation (including the self-defined occupation category ‘minor’), overnight travel in the past four weeks, nationality, and method of detection were calculated. Individual ages were rounded upward. For the analysis, samples were considered RDT-positive if they were positive for Pf only, or for both Pf and Pan. Demographic differences between all RDT-positive samples and those successfully sequenced, as well as between administrative regions, were assessed using Chi-square tests.

The proportion of samples carrying a given mutation or haplotype was calculated as the number of mixed and pure mutant genotypes divided by the total number of genotyped infections. The R function binom.test was used to estimate 95% confidence intervals for all prevalences.

The complexity of infection (COI) for each sample and population allele frequencies were jointly estimated for each municipality and province using MOIRE (v3.4.0), which implements a Markov chain Monte Carlo-based approach to jointly infer sample COI, within host relatedness, and population allele frequencies, using polyallelic genomic data.^54^ By modeling the genetic data and accounting for experimental errors, MOIRE provides probabilistic estimates. Microhaplotypes in all highly diverse *P. falciparum* targets (primer pool D1.1), as well as SNPs or microhaplotypes in drug resistance targets of interest, were used to infer COI and allele frequencies. The percentage of polyclonal infections was estimated as the mean of individual probabilities of polyclonality.

Univariate and multivariate logistic regression was performed to assess associations between the binary outcomes, presences of markers of interest and polyclonality (probability of polyclonality > 0.5), and key categorical variables, including age group, gender, occupation, detection method, and travel history. Analyses were restricted to mutations and variables with a sufficient sample size to achieve model convergence. Data from participants reporting travel to multiple countries were excluded from this analysis, as a single potential country of origin could not be assigned.

Non-*falciparum* species were considered present if the combined reads across all *ldh* targets exceeded 100, with each species contributing more than 1% of the total *ldh* target reads.

Putative *hrp2* and *hrp3* deletions were identified by applying a generalized additive model to account for target length amplification bias and differences in depth of coverage across primer pools as previously described.^52^ The model was fit on all data from the sequencing runs that included samples from this study, and also included samples from other countries in the GenE8 project that were sequenced in the same runs. To account for intrinsic target biases not explained by target length variation, a correction factor for targets of interest was obtained from the median residuals of all field samples and controls known not to have a deletion in each sequencing run. Read depth fold changes were estimated for *hrp2* and *hrp3*, and samples were labeled as *hpr2*- or *hrp3*-deleted if the estimated fold change was < 0.5. The workflow was validated using data from DBS controls made with cultured *P*. *falciparum* strain Dd2, which has a deletion in *hrp2* and was included in all sequencing runs.

To characterize genetic relatedness between samples, an infection-level relatedness estimate (r̂) based on identity-by-descent (IBD) was calculated for each sample pair using the *Dcifer* package and 165 highly heterozygous targets.^55^ The resulting r̂ value is an aggregate of the IBD between clones in the two infections in the analyzed pair. For this analysis, population allele frequencies were estimated by maximum likelihood using *Dcifer*’s *calcAFreq* function, and COI was estimated naively as the 95th quantile of the number of alleles observed at each target. Only samples with >75% of coverage across all targets were included in the analysis. Sample pairs were considered related if the null hypothesis that r̂ = 1 failed to be rejected at the 0.05 level. No correction was performed for multiple hypothesis testing because individual results are less of interest than the population-level patterns. Clusters of related samples were visualized by creating networks with samples as nodes and links drawn between nodes if the two samples were considered related, as defined above.

### Ethical Considerations

Ethics approval for the study was obtained from Eswatini Health and Human Research Review Board (EHHRRB013/2023) and the University of California San Francisco Institutional Review Board (350074). Written informed consent was obtained from all patients aged 18 years and older, as well as from parents/guardians of children aged 2 to 11 years. For patients aged 12 to 17 years, verbal assent, in addition to parent/guardian consent, was obtained. All participants or their parents/guardians were provided with a copy of the signed consent form. All personal identifiers were removed from survey data prior to storage on a password-protected cloud server, while the original signed consent forms were kept in a secure restricted-access location.

### Data availability

Deidentified individual-level genotypes, COI, and probability of polyclonality data can be found in the repository with the following DOI: 10.5281/zenodo.15387344.

## Results

### Sample characteristics

A total of 748 samples, comprising 713 DBS and 35 RDTs, along with the relevant survey data, were collected from 80 health facilities, beginning during the 2023 transmission peak (**Figure 1A-B**). Of these, 538 samples (504 DBS and 34 RDT) were collected from RDT-positive individuals, predominantly between April and June 2023 (**Figure S1A**). Most of the 210 RDT-negative samples were collected in the second half of 2023. Parasite density was estimated for 741 samples. Among PCR-confirmed RDT-positive samples, parasite density was higher in DBS extracts compared to RDT extracts (p<0.001, **Figure S1B**). Parasite densities ≥100 parasites/μL were observed in 83.4% of RDT-positive samples (439/526), in 5.4% of RDT-negative samples (11/204) and 2 of the 3 Pf-negative/Pan-positive samples (**Figure S1C**). The majority of RDT-negative (65.7%) had < 1 parasite/μL or undetectable parasite densities, with only 28.9% carrying parasite densities in the 1-100 parasite/μL range.

Sequencing libraries were prepared for 461 samples with parasite densities ≥100 parasites/μL and an additional 20 samples with parasite densities between 10 and 100 parasites/μL, representing 60 health facilities. The remaining health facilities did not have samples with parasite densities ≥100 parasites/μL. Potential cross-contamination led to the exclusion of 19 samples with parasite densities ≥100 parasites/μL and one sample with parasite densities between 10 and 100 parasites/μL. Four samples with <10,000 total reads were excluded. The median per-sample sequencing depth was 552,825 reads (IQR: 253,194-1,563,407) and the median per-target depth was 1,260 reads (IQR: 278-4,127). Sample sequencing depth was modestly correlated with parasite density (R^2^=0.2, p<0.001, **Figure S2A**). A median of 222 out of 238 targets per sample (93.3%, IQR: 198-225) had >100 reads. Markers with the lowest genotyping success rates included *crt* 218/220, *mdr1* 86 and PF3D7_1322700 236, which were successfully genotyped in 98, 351, and 357 of the 457 samples, respectively. Of the 457 samples analyzed, 392 (85.7%) had a genotype in all 59 *kelch13* SNPs assessed.

There were no significant demographic or epidemiological differences between all RDT-positive participants and those whose samples were successfully sequenced (**Table S1**). Most sequenced samples originated from the Hhohho (51%) and Lubombo (36%) regions (**Table 1**). Participants were predominantly male (78%) and aged between 15-39 years (68%). Nearly half (46%) of samples were collected from agricultural workers, who represented 63% of cases from the Hhohho region. Swazi nationals comprised the majority of participants with successfully sequenced samples (80%), followed by Mozambican nationals (20%). Most cases (93%) were detected through passive surveillance, and in the Hhohho region, 98% of cases were passively detected. Of all sequenced samples, 25% of the participants reported travel to Mozambique, and 7.5% reported travel within Eswatini. Compared to non-travelers, individuals who had traveled to Mozambique were more likely to be male, agricultural workers, and detected in Manzini region (**Table S2**).

**Table 1.** Characteristics of study participants with successfully sequenced samples.

|  | Overall | Hhohho | Lubombo | Manzini | Shiselweni |
| --- | --- | --- | --- | --- | --- |
| **Health facilities** | 54 | 17 | 22 | 12 | 3 |
| **N** | 392 | 200 | 140 | 45 | 7 |
| **Sex** |  |  |  |  |  |
| Female | 85 (22%) | 29 (15%) | 38 (27%) | 17 (38%) | 2 (29%) |
| Male | 305 (78%) | 171 (86%) | 102 (73%) | 28 (62%) | 5 (71%) |
| **Age** |  |  |  |  |  |
| <5 | 12 (3.1%) | 3 (1.5%) | 7 (5.1%) | 2 (4.5%) | 0 (0%) |
| 5-14 | 50 (13%) | 19 (9.5%) | 22 (16%) | 7 (16%) | 2 (29%) |
| 15-24 | 134 (35%) | 74 (37%) | 46 (34%) | 11 (25%) | 3 (43%) |
| 25-39 | 127 (33%) | 81 (41%) | 40 (29%) | 6 (14%) | 0 (0%) |
| 40-59 | 48 (12%) | 17 (8.5%) | 16 (12%) | 13 (30%) | 2 (29%) |
| >59 | 16 (4.1%) | 5 (2.5%) | 6 (4.4%) | 5 (12%) | 0 (0%) |
| Unknown | 5 | 1 | 3 | 1 | 0 |
| **Occupation** |  |  |  |  |  |
| Minor | 16 (4.1%) | 4 (2.0%) | 9 (6.5%) | 3 (6.8%) | 0 (0%) |
| Student | 84 (22%) | 30 (15%) | 40 (29%) | 10 (23%) | 4 (57%) |
| Agricultural | 179 (46%) | 125 (63%) | 51 (37%) | 3 (6.8%) | 0 (0%) |
| Other | 41 (11%) | 15 (7.5%) | 6 (4.3%) | 18 (41%) | 2 (29%) |
| Unemployed | 69 (18%) | 26 (13%) | 32 (23%) | 10 (23%) | 1 (14%) |
| Unknown | 3 | 0 | 2 | 1 | 0 |
| **Nationality** |  |  |  |  |  |
| Eswatini | 312 (80%) | 157 (79%) | 122 (88%) | 27 (61%) | 6 (86%) |
| Mozambique | 78 (20%) | 43 (22%) | 17 (12%) | 17 (39%) | 1 (14%) |
| Unknown | 2 | 0 | 1 | 1 | 0 |
| **Travel** |  |  |  |  |  |
| No Travel | 253 (65%) | 140 (70%) | 94 (67%) | 15 (34%) | 4 (57%) |
| Domestic | 29 (7.4%) | 7 (3.5%) | 16 (11%) | 6 (14%) | 0 (0%) |
| Mozambique | 98 (25%) | 49 (25%) | 27 (19%) | 21 (48%) | 1 (14%) |
| Other countries | 9 (2.3%) | 2 (1.0%) | 3 (2.1%) | 2 (4.5%) | 2 (29%) |
| Multiple countries | 2 (0.5%) | 2 (1.0%) | 0 (0%) | 0 (0%) | 0 (0%) |
| Unknown | 1 | 0 | 0 | 1 | 0 |
| **Detection method** |  |  |  |  |  |
| Passive | 364 (93%) | 196 (98%) | 119 (86%) | 43 (98%) | 6 (86%) |
| Reactive | 24 (6.2%) | 4 (2.0%) | 18 (13%) | 1 (2.3%) | 1 (14%) |
| Active | 2 (0.5%) | 0 (0%) | 2 (1.4%) | 0 (0%) | 0 (0%) |
| Unknown | 2 | 0 | 1 | 1 | 0 |

### Genomics reveals low levels of antimalarial drug resistance markers of concern

Among the 443 samples with adequate sequencing depth for *hrp2*/*3* deletion analysis, only 3 were from RDT-negative individuals. No evidence of *hrp2*/*3* gene deletions was detected in any sample. Non-*falciparum* co-infections were rare: *P*. *malariae* was detected in only 3 samples (0.7%), 2 from the Lubombo region and one from the Hhohho region. These included one Swazi and 2 Mozambican nationals who reported travel to Mozambique. No co-infections with *P*. *ovale* or *P*. *vivax* were observed.

Overall, 61.4% of samples (95% confidence interval, CI: 59.9-63.2%) were polyclonal, with a mean COI of 2.54 (95% CI: 2.48-2.61). The proportion of polyclonal infections and mean COI were significantly lower in the Hhohho region compared to the other three regions (**Table 2**). Participants who reported travel to Mozambique were twice as likely to carry polyclonal infections (odds ratio [OR] 2.51, 95% CI: 1.44-4.44, **Table S3**). In contrast, samples with higher parasitemia (OR: 0.71, 95% CI: 0.59-0.86, **Figure S2B**) and higher sequencing depth (OR: 0.58, 95% CI: 0.38-0.86) were less likely to be polyclonal. No significant associations were observed between polyclonality and participant demographics. Mixed genotypes in at least one drug resistance marker were detected in 39.3% of the samples.

**Table 2.** Complexity of infection (COI) and percentage of polyclonal infections for each administrative region in Eswatini. Confidence intervals (CI) were drawn from posterior distributions.

|  | Overall | Hhohho | Lubombo | Manzini | Shiselweni |
| --- | --- | --- | --- | --- | --- |
| *N* | 456 | 238 | 162 | 49 | 7 |
| Mean COI<br>[95% CI] | 2.54<br>[2.48-2.61] | 2.37<br>[2.29-2.45] | 2.72<br>[2.62-2.82] | 2.67<br>[2.51-2.88] | 3.61<br>[3.14-4.42] |
| Percentage<br>polyclonal | 61.4%<br>[59.9-63.2%] | 55.6%<br>[53.4-58.4%] | 65.1%<br>[61.1-68.5%] | 74.1%<br>[69.4-77.6%] | 85.4%<br>[71.4-100%] |
| [95% CI] |  |  |  |  |  |

### Mutations in dhps and dhfr

Mutations in the *dhps* and *dhfr* genes - associated with resistance to sulfadoxine and pyrimethamine, respectively - were highly prevalent (Table S4). The *dhps* ISGEAA double mutant haplotype (mutations A437G and K540E; wild type at I431, S436, A581, and A613) was detected in 94.4% of the samples (N=429), with the highest prevalence in the Hhohho region (96.4%, N=225, **Table S4**). The ISGEGA triple mutant haplotype (carrying an additional A613G mutation) was observed in 6.8% of samples overall, reaching 12.8% (N=47) in the Manzini region. Wild type (*dhps* ISAKAA) and the I431V-containing variant (VAGKGS) were rare, each observed in <3% of samples. No triple *dhps* mutant haplotypes ISGEGS or ISGEAS were detected. Haplotype inference was not possible for 22 samples (5.1%). The *dhfr* IRNI triple mutant haplotype (mutations N51I, C59R, S108N; wild type I164) was nearly fixed, present in 98.9% of samples (N=453, **Table S4**). A single sample (0.2%) carried the wild-type NCSI haplotype, with haplotype inference not possible for one sample. The I164L mutation was not detected in any of the samples analyzed. Among the 429 samples with complete *dhps*/*dhfr* haplotypes (**Table 3**), 93.2% carried the quintuple mutant haplotype (*dhps* ISGEAA and *dhfr* IRNI), while 6.8% carried the sextuple mutant haplotype (*dhps* ISGEGA and *dhfr* IRNI). All non-ISGEAA *dhps* haplotypes were found in combination with the *dhfr* IRNI haplotype.

**Table 3.**
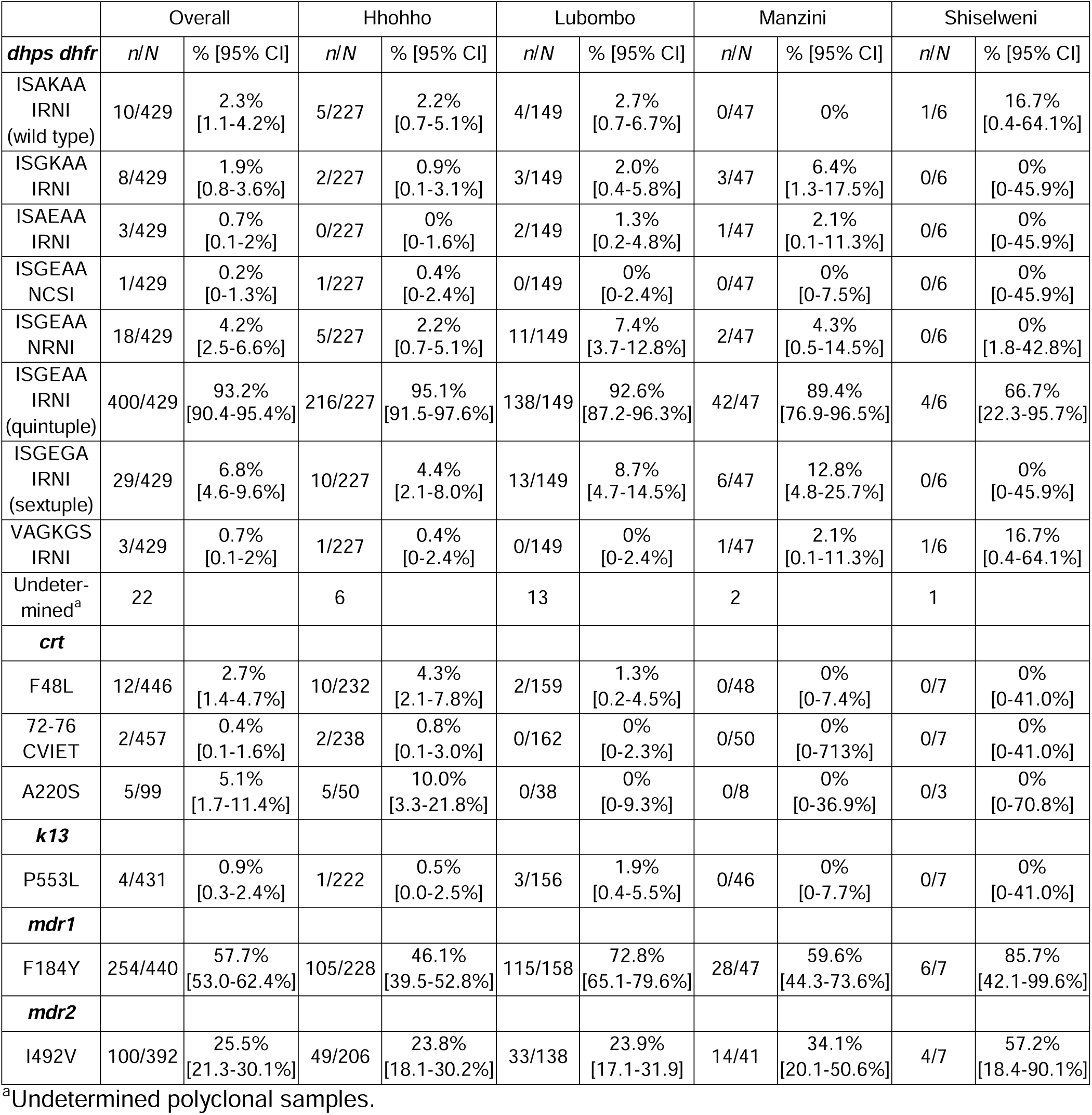
Proportion of samples carrying antimalarial drug resistance markers in Eswatini.

Multivariate analysis indicated that participants reporting recent international travel to countries other than Mozambique were less likely to carry the quintuple mutant haplotype, though the sample size was small (OR: 0.05, 95% CI: 0.01-0.23, **Table S5**). All infections carrying the sextuple mutant haplotype were detected through passive surveillance between April and May 2023 (**Figure 2**), coinciding with the peak in sample collection (**Figure 1C**). No association with gender, age, occupation, or parasitemia, was found for either the quintuple or the sextuple mutant haplotypes. The 3 samples carrying the *dhps* VAGKGS haplotype were monoclonal and geographically dispersed. Of those, 2 were collected in the Hhohho and Shiselweni regions from patients reporting recent travel to Tanzania. Both presented to health facilities within 3 days of each other in July 2023 (**Figure 2**). The third, from Manzini, came from a participant with no reported travel and presented to the health facility 44 days later.

**Figure 2.**
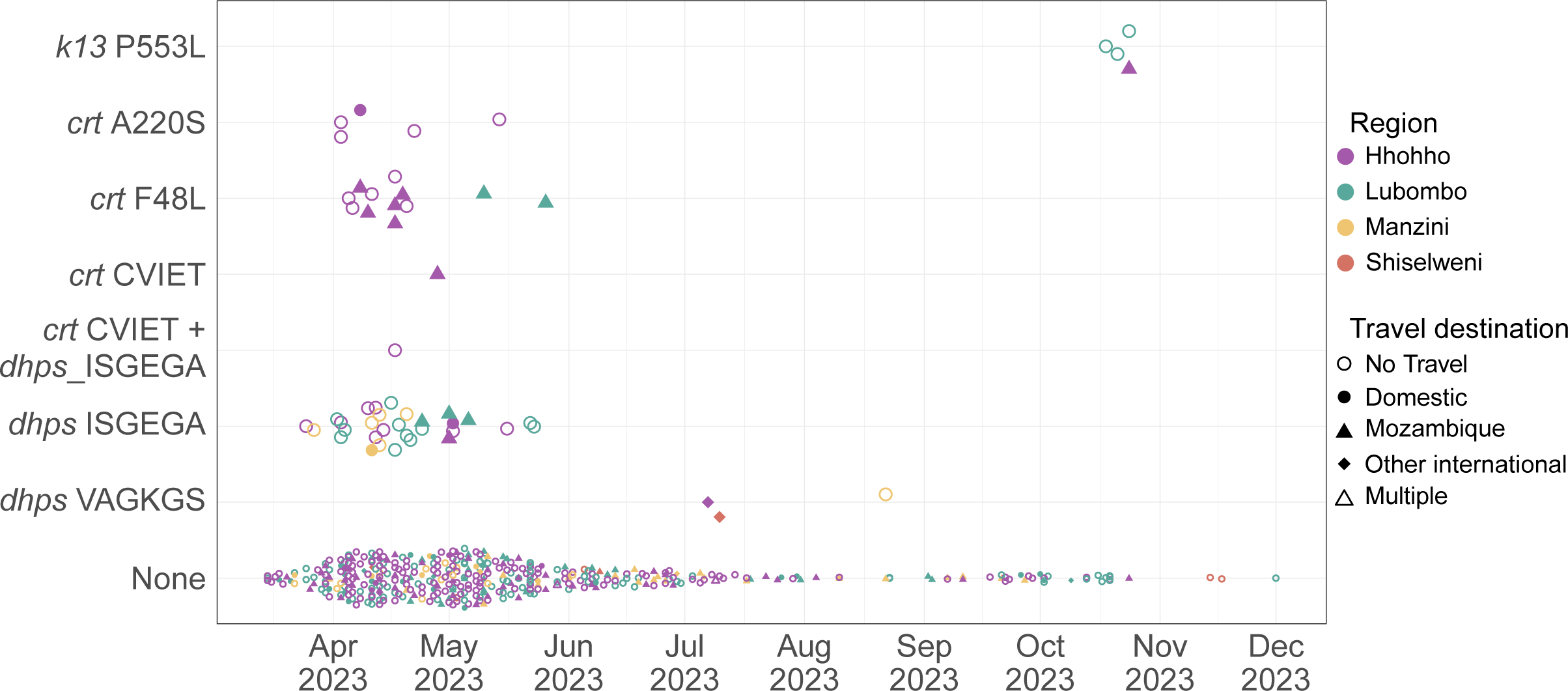
Temporal distribution of samples carrying genotypes of interest. Collection date and region, genotype and travel history for each sample in the dataset.

### Mutations in crt

The *crt 72-76 CVIET* mutant haplotype - associated with chloroquine resistance - was detected in only 2 of 457 samples (0.4%, 95% CI: 0.1-1.6%), both from the Hhohho region. These samples were polyclonal and carried mixed *crt* 72-76 mutant CVIET and wild-type CVMNK haplotypes. The *crt* A220S mutation was found in 5 of 99 samples with genotype data for this locus (5.1%, 95% CI: 1.7-11.4%), all from the Hhohho region, representing 10% of samples sequenced in that region. All A220S mutations co-occurred with the wild-type *crt* 72-76 CVMNK haplotype. The *crt* F48L mutation was detected in 12 samples (2.7%, 95% CI: 1.4-4.7%), including 10 from Hhohho (4.3%, 95% CI: 2.1-7.9%) and 2 from Lubombo (1.3%, 95%CI: 0.2-4.5%). These also occurred exclusively with the wild-type *crt* 72-76 CVMNK haplotype. All *crt* mutations were found in samples collected between April and May 2025, in both travelers and non-travelers (**Figure 2**). The *crt* I356T mutation was not observed.

### Mutations in kelch13

The only *kelch13* propeller domain mutation identified was P553L, a validated marker of artemisinin partial resistance. It was detected in 4 of 456 samples (0.9%, 95% CI: 0.3-2.4%): 1 from Hhohho (0.5%, 95%CI: 0.0-2.5%) and 3 from Lubombo (1.9%, 95%CI: 0.4-5.5%). All four samples were collected in the second half of October 2023 (**Figure 2**), with 3 of the 4 samples polyclonal. One case, from Hhohho, involved a traveler to Mozambique; the rest reported no travel. Of the P553L-positive individuals, 3 were agricultural workers and one was a minor.

### Mutations in mdr1 and mdr2

The *mdr1* N86 wild-type genotype was found in all 351 samples with a valid genotype in that locus. No samples carried the *mdr1* N86Y genotype. In contrast, the *mdr1* F184Y genotype was detected in 57.6% of the samples (95% CI: 52.9-62.3%), with regional prevalence ranging from 39.5% in Hhohho (95%CI: 39.5-52.8%) to 85.7% in Shiselweni (95% CI: 42.1-99.6%, **Table 3**). This mutation was not significantly associated with patient characteristics (**Table S7**). The *mdr2* I492V mutation was identified in 25.6% of the samples (95% CI: 21.3-30.2%), with similar prevalence across all 4 regions (**Table 3**). Like *mdr1* F184Y, this mutation was not significantly associated with demographic or clinical variables (**Table S8**).

### Samples carrying mutations form clusters of highly related infections

The IBD-based measure of infection-level genetic relatedness (r̂) among samples was bimodal, with peaks at 0 and 1 (**Figure S3**). Using a threshold of r̂ = 1 as the null hypothesis, 235 of 405 samples (58.0%) were classified as highly related to at least one other sample, forming 988 highly related sample pairs (1.21% of all sample pairs). Clustering analysis revealed widespread genetic clustering: 6 clusters contained more than 5 samples, while 36 additional clusters consisted of 5 or fewer samples (**Figure 3A**). Lowering the relatedness threshold to r̂ > 0.5 increased the size of the clusters but did not change overall cluster patterns (**Figure S4A**). Polyclonality was significantly more frequent in samples not assigned to any cluster (61.8%) compared to those within clusters (41.3%, p<0.001, chi-squared test). Similarly, unrelated samples were significantly more likely to come from participants reporting international travel (41.3% vs. 17.2%, p<0.001, chi-squared test). Clusters were typically composed of samples collected from the same region and similar time frames, although some spanned multiple months and regions (**Figure 3B**).

**Figure 3.**
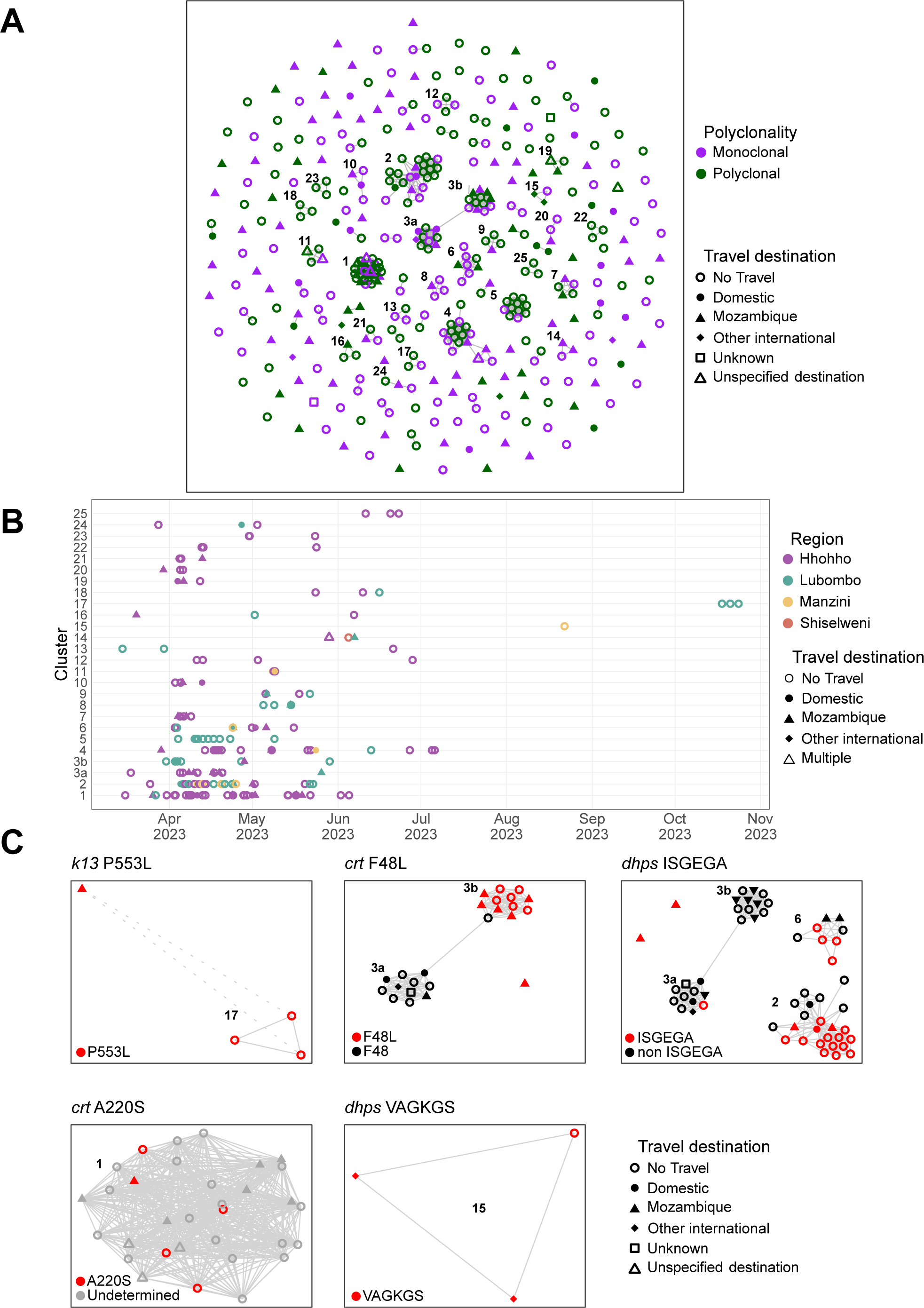
Genotypes of interest are segregated in genetically related clusters. **A**. Clusters of genetically related samples, defined as sample pairs with a relatedness value of r̂ =1, at the 0.05 significance level. Only clusters composed of 3 or more samples are labeled in the top-left of the cluster. **B**. Samples belonging to each cluster of 3 or more samples by collection date and region. **C**. All clusters and isolated samples containing a genotype of interest are shown indicating the genotype for each sample in those clusters. Cluster labels are the same as panels A and B. Missing genotypes due to low depth of coverage are indicated in grey. Dashed lines indicate significantly related pairs with a lower r̂ (0.5) only shown for samples carrying *kelch13* P553L.

Although mutations in the *dhps* (ISGEGA and VAGKGS haplotypes), *crt* (F48L, 72-76 CVIET and A220S), and *kelch13* (P553L) genes were detected at low prevalence, they were often observed in temporally clustered samples and, in some cases, within close geographic proximity. These mutations of interest were mostly segregated within related clusters (**Figure 3C**). Of the 4 samples carrying *kelch13* P553L, 3 were part of cluster 17, while the fourth showed significant relatedness to samples in that cluster when the threshold used for hypothesis testing was lowered to 0.5 (**Figure S4B**). All but one sample carrying the *crt* F48L mutation belonged to cluster 3, which contained 23 samples organized into 2 tightly connected subclusters (3a and 3b), overlapping in time and linked by polyclonal infections. The F48L mutation was restricted to subcluster 3b. These subclusters included samples from multiple individuals who had traveled to Mozambique. All samples with a valid A220S mutation were detected exclusively in cluster 1. All other samples in this cluster lacked genotype calls for amino acid 220, and none were successfully called as wild type. Cluster 1 was predominantly composed of samples collected in the Hhohho region, with several from participants reporting travel to Mozambique. All 3 samples carrying the *dhps* VAGKGS haplotype were found in cluster 15. The *dhps* ISGEGA haplotype was distributed across 3 mixed *dhps* genotype clusters (clusters 2, 3a, and 6), and also appeared in 2 unclustered samples from travelers to Mozambique. Cluster 2 included the majority of samples with *dhps* ISGEGA, which included several tightly-connected samples from non-travelers as well as 2 from travelers to Mozambique. These mutations (*dhps* ISGEGA or VAGKGS; *crt* F48L, CVIET or A220S; or *k13* P553L) were present in 11.9% of all samples and were more common in larger clusters. While 25.2% of all samples were part of clusters with more than 5 samples, 58.3% of samples carrying at least one of these mutations belonged to such clusters.

## Discussion

This malaria molecular surveillance study, leveraging targeted amplicon sequencing, provides the first data on antimalarial drug and diagnostic resistance markers from Eswatini since 2010. In addition to assessing marker prevalence, parasite genomic data were used to characterize population structure and transmission dynamics, offering critical insights into the emergence and spread of drug resistance parasites in low endemic settings. The validated ART-R marker, *kelch13* P553L, was detected at low prevalence but formed a genetically distinct cluster of highly related infections in the Hhohho and Lubombo regions. Although the *crt* K76T chloroquine resistance marker was nearly absent, sulfadoxine-pyrimethamine (SP) resistance markers were nearly fixed: 95% of samples carried the quintuple mutant haplotype (*dhps* ISGEAA + *dhfr* IRNI), and 7% carried the sextuple mutant haplotype (*dhps* ISGEGA + *dhfr* IRNI). These haplotypes were concentrated in transmission clusters, supporting the role of local amplification. The *mdr1* N86 wild-type genotype - associated with lumefantrine tolerance - was present in all samples.^19^ Non-*falciparum* species were very rare, and no *hrp2*/*3* deletions were identified.

Eswatini relies exclusively on artemether-lumefantrine for the treatment of uncomplicated malaria. The rapid emergence and spread of ART-R in East and the Horn of Africa, as well as recent reports of reduced lumefantrine susceptibility and decreased artemether-lumefantrine efficacy, pose a growing risk of de novo emergence or introduction of ART-R in Eswatini.^6,9,56,57^ The detection of the validated ART-R marker *kelch13* P553L in a localized cluster - one case linked to travel from Mozambique and others with no travel history - raises concern for both importation and onward transmission. This mutation has been associated with delayed clearance in Southeast Asia but has been rarely reported in Sub-Saharan Africa.^12,58–60^ Notably, this same mutation was recently reported in South Africa’s Mpumalanga province, which borders Eswatini to the north and west, in October 2023.^61^ Whether parasites carrying this mutation persisted into the 2023/2024 transmission season remains unknown. Additional research is of critical importance to assess its clinical impact in southern Africa and to determine the ancestral origin of the parasites identified in this dataset. No other *kelch13* mutations identified in other African countries, were detected in this study.^6,53,62–64^

Even in the presence of ART-R, ACTs are expected to remain effective due to the action of the partner drugs. Although reduced lumefantrine susceptibility has been reported in Uganda, no full resistance has been documented, and no validated genetic markers of lumefantrine resistance currently exist.^6,56^ The *mdr1* N86 genotype, associated with a modest reduction in susceptibility in laboratory studies and shown to be selected for by artemether-lumefantrine use, was found in all samples in this study, possibly reflecting the selective pressure of the drug.^7,56,65^ Molecular markers of resistance to other ACT partner drugs, such as mutations in *crt* and *mdr1* associated with piperaquine and amodiaquine resistance, were not detected.^18,19^ Duplications of *mdr1* or *plasmepsin 2*/*3* linked to mefloquine and piperaquine resistance, respectively, were not assessed.^20,21^

Mutations conferring resistance to former first-line antimalarials, chloroquine and SP, showed patterns consistent with current drug use and malaria epidemiology in the region. The chloroquine resistance-associated *crt* CVIET haplotype, previously widespread under chloroquine pressure, was nearly absent.^36^ The low-prevalence *crt* A220S and F48L mutations require further investigation. A220S can confer higher levels of chloroquine resistance in the presence of the *crt* CVIET haplotype.^66^ In this study, only a fourth of the samples had a valid genotype at *crt* 220 due to low amplification efficiency, and all samples with A220S were found in conjunction with the wild-type *crt* CVMNK haplotype. The *crt* F48L mutation has not been linked to resistance to any antimalarials, but it has been detected at very low levels in other countries in southern Africa.^62,63^ SP has not been used for treatment in Eswatini since the adoption of artemether-lumefantrine in 2010, but remains widely used for chemoprophylaxis across Sub-Saharan Africa.^67^ The high prevalence of the *dhps*/*dhfr* quintuple mutant haplotype in Eswatini likely reflects importation from Mozambique, where SP is routinely used in intermittent preventive treatment in pregnancy.^68^ The sextuple mutant haplotype, associated with higher SP resistance, was found in 7% of samples, higher than the prevalence recently reported in Mozambique and South Africa (<1%).^61,68^ A cluster of three samples carrying the *dhps* VAGKS haplotype was detected. This haplotype has been mainly reported in West and Central Africa and seems to be selected for by SP use in pregnancy.^69,70^

Genetic relatedness analysis revealed substantial clustering among infections, with a large proportion of samples belonging to highly related clusters, which frequently included samples carrying key resistance markers. Over 70% of the cases reported no recent international travel, highlighting the risk for local amplification of resistant strains in low transmission settings such as Eswatini. The generated genetic data combined with travel histories for samples carrying the *kelch13* P553L mutation - a validated marker of ART-R - do not clarify the linkage between these cases. The infection in the traveler to Mozambique could have originated the three locally acquired cases, if the traveler had remained gametocyte-positive during the month between arrival and sample collection, or the parasite may have been related to a missed common ancestor of the locally acquired cases. De novo emergence of the mutation within Eswatini cannot be ruled out. Similarly, the *dhps* VAGKGS haplotype appears to have been co-imported by two travelers from Tanzania, linked to a single secondary infection detected six weeks later. In contrast, clusters containing *crt* F48L or A220S mutations included separate travelers to Mozambique suggesting that multiple introduction events may have seeded these clusters. Samples carrying the sextuple mutant haplotype ISGEGA were found in three large clusters, each of which included at least one traveler to Mozambique. These patterns suggest that the rapid spread of parasites contributed to the unexpectedly high prevalence of this marker in Eswatini. Overall, these results highlight how cross-border importation and local amplification can be interlinked processes that shape the epidemiology of antimalarial drug-resistance, even in the presumptive absence of local drug selective pressure. Rapid expansion of highly related parasite populations may facilitate the emergence of resistant genotypes in low transmission settings. Further research is needed to infer fine-scale transmission networks to clarify pathways of parasite spread.

Higher malaria transmission intensity is typically associated with greater COI.^39,71,72^ Over 60% of the samples in this study were polyclonal. Although polyclonality was more common among travelers to Mozambique, likely reflecting the higher parasite diversity in source populations, more than half of infections among individuals who reported no international travel were also polyclonal. This suggests that ongoing local transmission is contributing to superinfections and cotransmission of multiple clones within transmission chains. The negative association between parasite density and COI may reflect host immune responses in a population with low immunity interacting with within-host parasite diversity and/or intra-host parasite interactions, and warrants further investigation.

A previous study in Eswatini, conducted on patients presenting to health facilities, demonstrated that of 162 PCR-detectable infections, 33% were subpatent and reported no *hrp2/3* deletions.^32^ The majority of RDT-negative samples analyzed in this study (both Pf- and Pan-negative) were PCR-negative or had very low parasite density, and 28.1% had parasite densities in the 1-100 parasites/μL range. Of the RDT-negative samples, 5% had parasite densities above 100 parasites/μL, which could be due to mislabeling, database errors, or the prozone-like effect. HRP2 antigen blood concentration was not measured in these samples. Among the RDT-negative and Pf-negative/Pan-positive samples with > 100 parasites/μL, only a subset had sufficient sequencing depth to assess for *hrp2*/*3* deletions, but no putative *hrp2*/*3* deletions were identified in either RDT-negative or RDT-positive samples. These findings, along with recent data from Mozambique and South Africa, suggest that there is no evidence for RDT misdiagnosis due to *hrp2*/*3* deletions.^61,73^ A recent study from South Africás KwaZulu-Natal province, which borders Eswatini’s Hhohho and Shiselweni regions, reported a high prevalence of *hrp2*/*3* deletions using nested PCR, but the small sample size, together with methodological and sample quality differences, may explain the discrepancy.^74^ Overall, our data confirms subpatent infections occur even in symptomatic cases identified at health facilities.

This study had several limitations. Sample collection began at the peak of the 2022-2023 transmission season, missing the critical early-season travel period in December and January. As a result, we were unable to capture early-season transmission dynamics and may have introduced bias into prevalence estimates due to the temporal and spatial clustering of cases. Additionally, RDT-negative individuals were sampled in the second half of 2023, when malaria transmission and test positivity rates are typically lowest, further reducing our ability to detect and quantify *hrp2/3* deletions. Finally, we acknowledge that asymptomatic infections and those occurring in harder-to-reach populations are less likely to be captured in this dataset due to data collection through passive surveillance systems. Although we targeted all malaria cases within a single transmission season, the achieved sample size was smaller than anticipated due to the delayed start, limiting both the precision of prevalence estimates and the statistical power to detect associations between mutations and case characteristics. Approximately 15% of RDT-positive samples yielded incomplete sequencing results, although these were evenly distributed across time and region, and no socio-demographics differences were observed, reducing the likelihood of selection bias. Technical limitations of the sequencing assay further impacted data completeness. Uneven read depth across loci reduced the effective sample size for certain markers, particularly *crt* mutations. Additionally, phasing SNPs across amplicons to reconstruct haplotypes was not always possible.

The revolution in genomic technologies has opened new avenues to study adaptation and spread of malaria parasites within and across borders, offering valuable epidemiological insights for improving elimination strategies. This genomic surveillance study identified clusters of genetically related infections carrying markers of antimalarial drug resistance, including ART-R. While overall prevalence of these markers was low, the findings support a pattern of introduction and local spread as the mechanism for resistance emergence in low transmission settings. Therefore, routine molecular surveillance to track trends and deliver timely data is urgently needed to inform interventions to prevent the spread of resistant parasites in low transmission areas.

## Acknowledgements

We thank the participants, nurses, field supervisors and data managers that made this study possible. We thank the colleagues from the National Malaria Programme for their support and collaboration throughout. We also thank Hazel Gwarinda, Nokwethemba Kubheka, Ongeziwe Taku from the NICD for their laboratory support, as well as Nick Hathaway and Sophie Bérubé for technical assistance. We also thank members of the Malaria Elimination Initiative, the EPPIcenter and the IDDynamics group at JHU for productive discussions.

