## Supplementary Material for "Genomic Surveillance Reveals Clusters of *Plasmodium falciparum* Antimalarial Resistance Markers in Eswatini, a Low-Transmission Setting"

**List of supplementary figures and tables:**

**Figure S1.** Sample RDT results and parasite density.

**Figure S2.** Relationship between parasite density, depth of coverage, and intrahost diversity.

**Figure S3:** Distribution of estimated relatedness ($\hat{r}$) between all sample pairs.

**Figure S4:** Clustering of samples with a relatedness threshold of 0.5.

**Table S1.** Comparison of characteristics and origin of recruited RDT-positive participants and those with successfully sequenced samples.

**Table S2.** Comparison of characteristics and origin of recruited participants who traveled to Mozambique to those who did not travel or traveled to other countries.

**Table S3.** Logistic regression of infection polyclonality and case characteristics.

**Table S4.** Proportion of samples carrying a *dhps* or *dhfr* haplotype.

**Table S5.** Logistic regression of *dhps*/*dhfr* quintuple mutant carriage and case characteristics.

**Table S6.** Logistic regression of *dhps*/*dhfr* sextuple mutant carriage and case characteristics.

**Table S7.** Logistic regression of *mdr1* F184Y and case characteristics.

**Table S8.** Logistic regression of *mdr2* I492V and case characteristics.

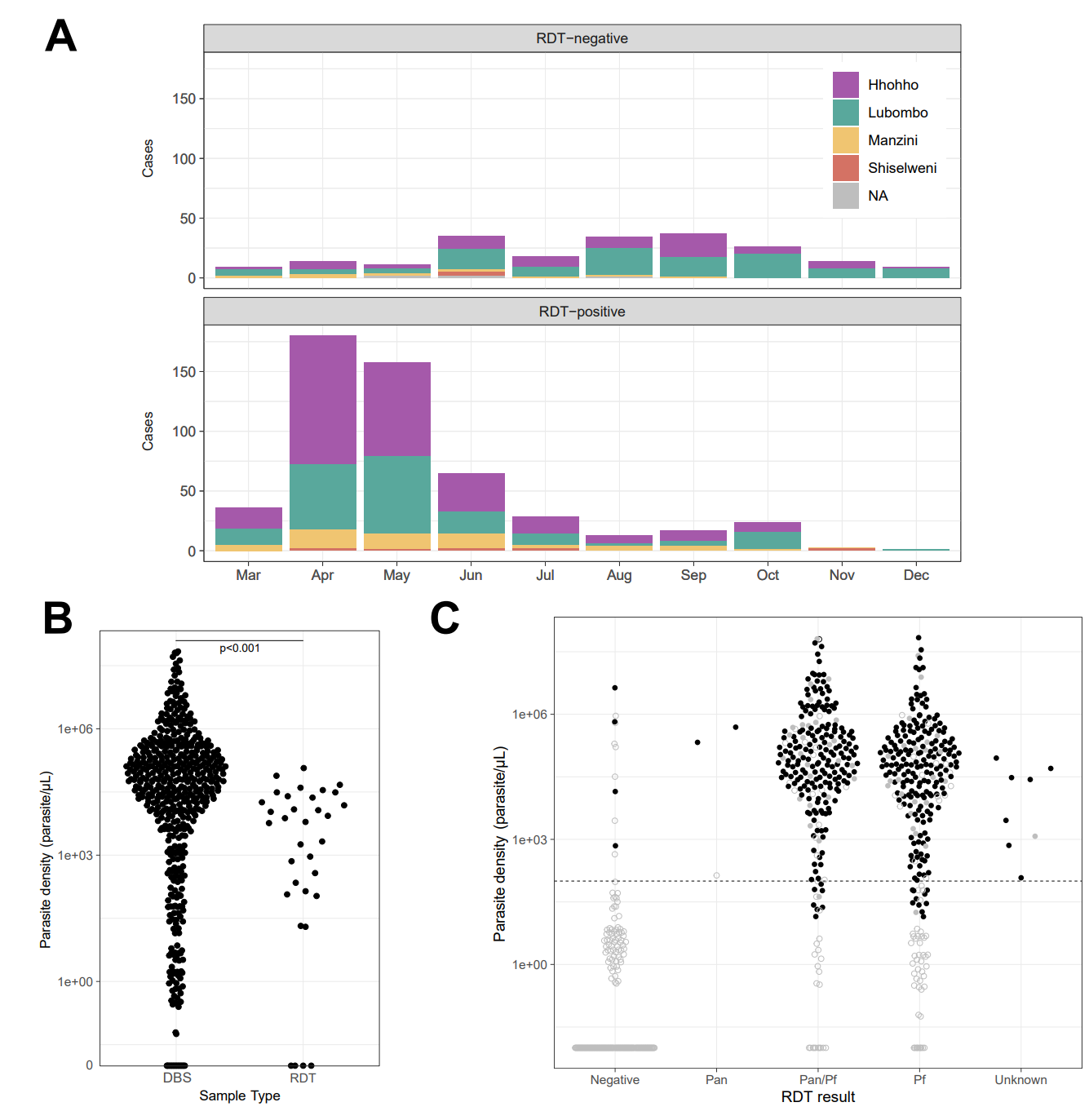

**Figure S1. Sample RDT results and parasite density. A.** Time of collection of RDT-positive (Pf or Pf/Pan RDT result) or RDT-negative (Negative or Pan-only) samples. **B.** Difference in parasite density in DNA extracts from DBS or RDT from RDT-positive samples. The reported p-value corresponds to a Wilcoxon rank-sum test. **C.** Parasite density in samples with different reported RDT results. Samples with a genotype in all 59 assessed kelch13 SNPs are indicated in closed circles, and those missing at least one SNP in open circles. Samples with a valid *hrp2*/*3* target depth fold change estimate are indicated in black, and those withour sufficient depth of coverage in grey. None of the samples with a valid *hrp2*/*3* target depth fold change were classified as *hrp2*- or *hrp3*-deleted. The dashed line indicates 100 parasites/μL.

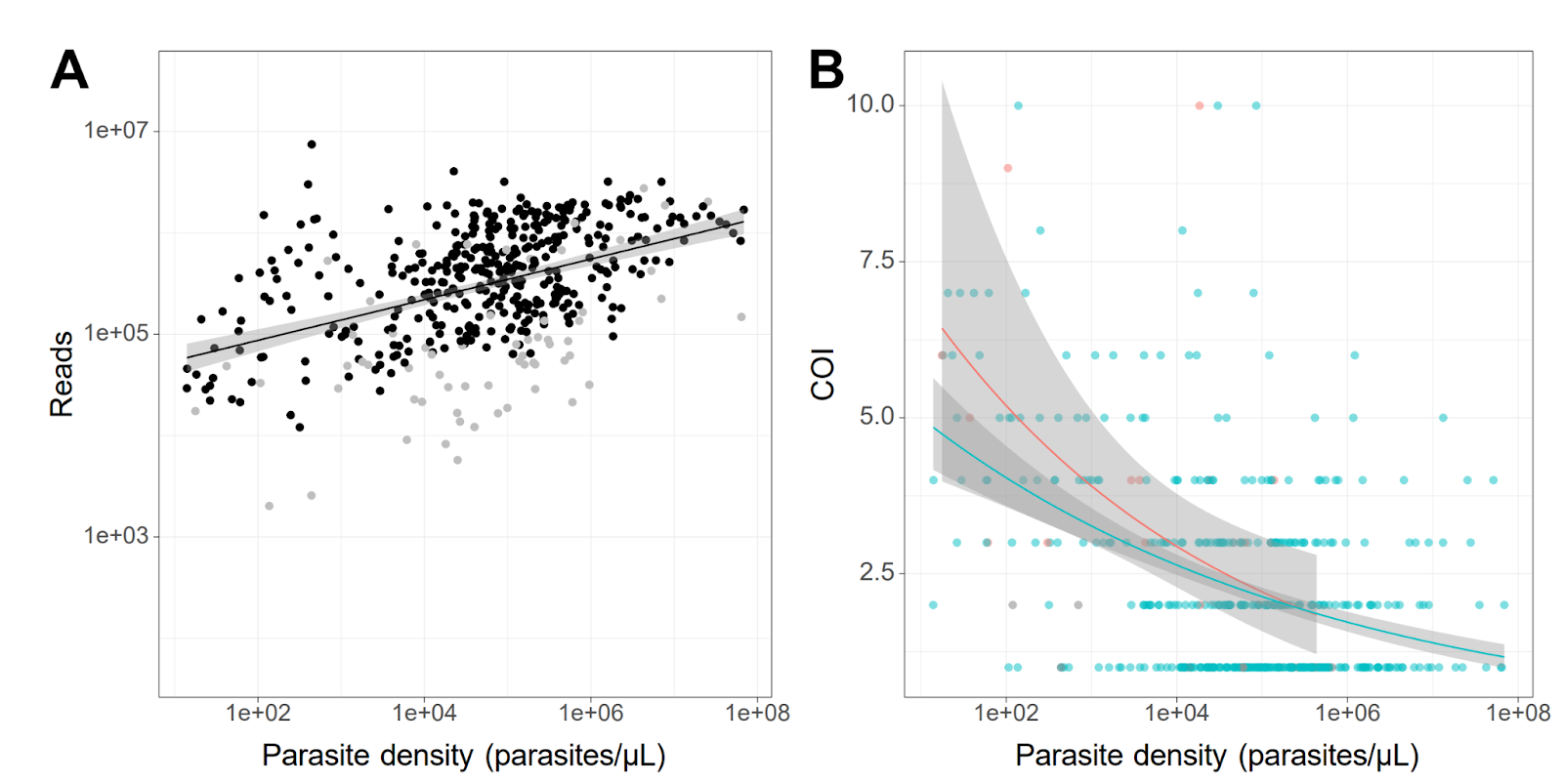

**Figure S2. Relationship between parasite density, depth of coverage, and intrahost diversity. A.** Linear regression of sample parasite density and total sample reads, for samples with a valid genotype in all 59 assessed *kelch13* SNPs (black) and those missing at least 1 valid SNP (grey). R^2^=0.19, p<0.001. **B.** Quasi-Poisson regression of sample parasite density and complexity of infection (COI) for samples collected passively (cyan) or by proactive and reactive case detection (red). Grey samples are RDT-negative.

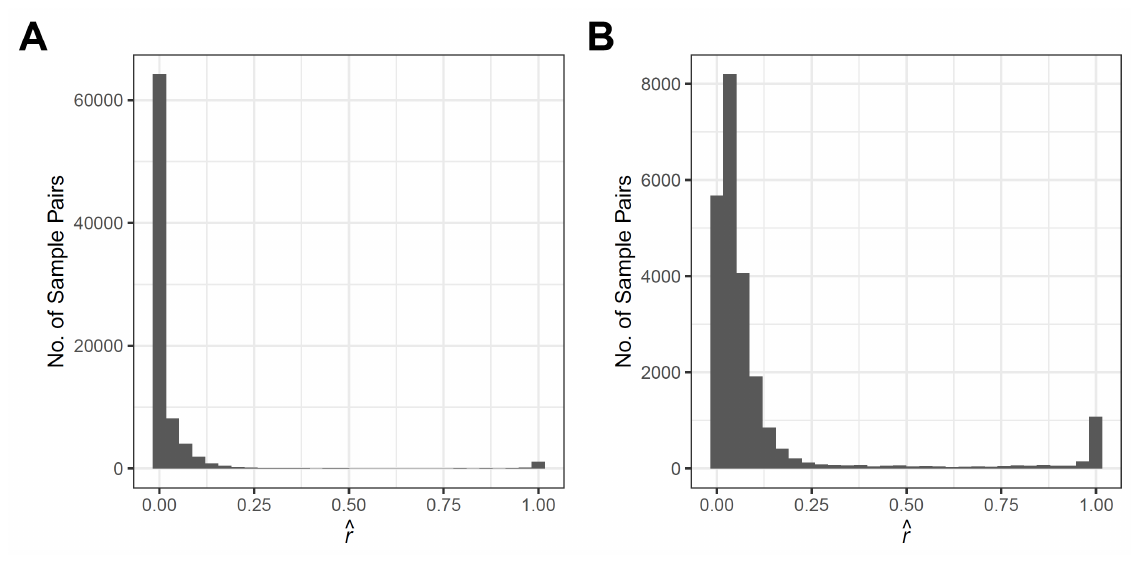
**Figure S3: Distribution of estimated relatedness (**$\hat{\boldsymbol{r}}$**) between all sample pairs. A.** Histogram showing $\hat{r}$ among all pairs of samples in the dataset. **B.** As A, except pairs with an $\hat{r}$ of zero have been removed, to better visualize the remainder.

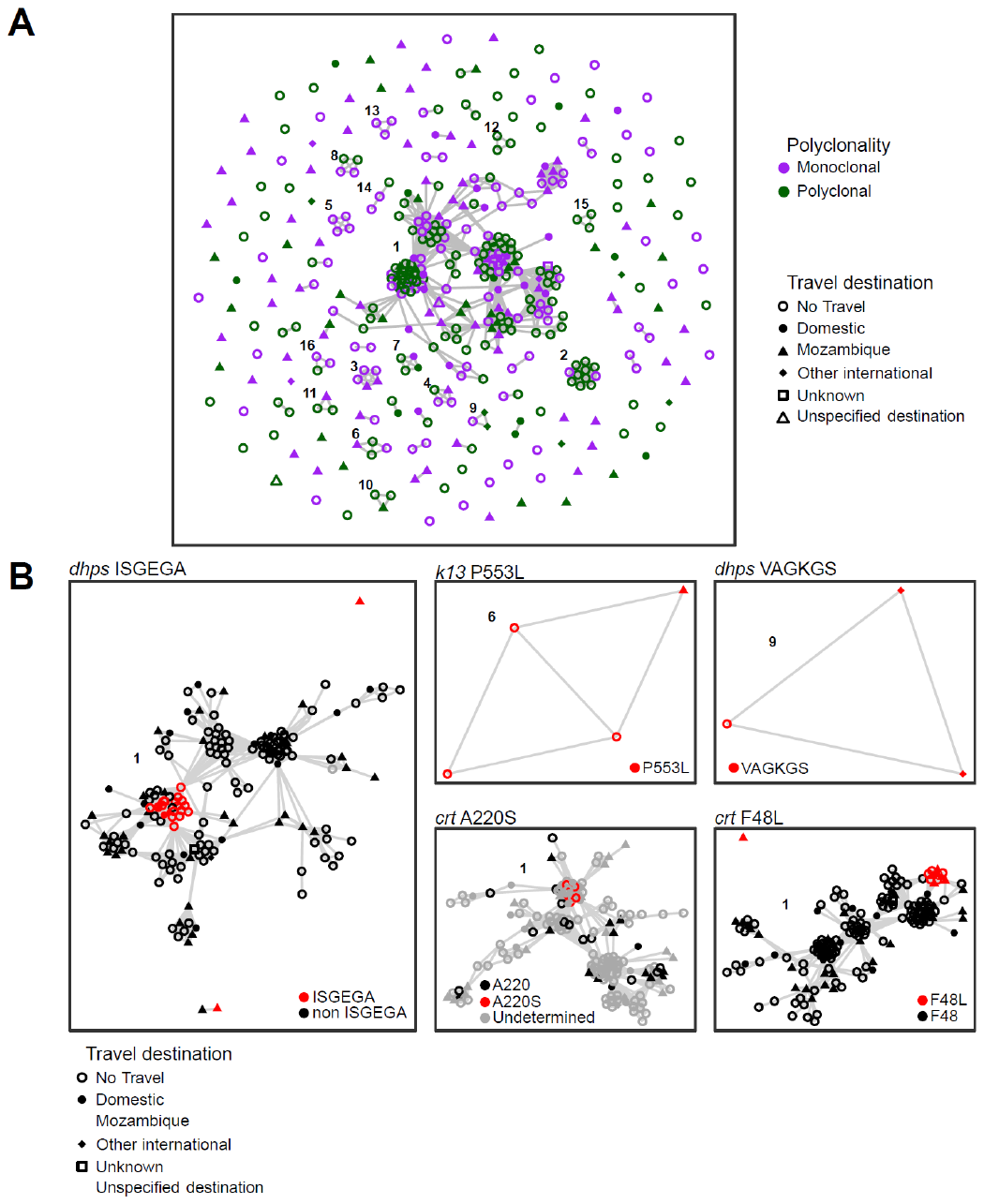

**Figure S4: Clustering of samples with a relatedness threshold of 0.5. A.** Clusters of genetically related samples, defined as sample pairs with a relatedness value of $\hat{r}$>0.5, at the 0.05 significance level. Only clusters composed of 3 or more samples are labeled in the top-left of the cluster. **B.** All clusters and isolated samples containing a genotype of interest are shown indicating the genotype for each sample in those clusters. Cluster labels are the same as panel A.

**Table S1. Comparison of characteristics and origin of recruited RDT-positive participants and those with successfully sequenced samples**

|  | **RDT-positive** | **Successfully sequenced*** | ***P*-value**** |
| --- | --- | --- | --- |
| ***N*** | 572 | 379 |  |
| **Sex** |  |  | 0.06 |
| Female | 156 (27%) | 82 (22%) |  |
| Male | 415 (73%) | 297 (78%) |  |
| **Age** |  |  |  |
| <5 | 16 (2.8%) | 11 (2.9%) | 0.97 |
| 5-14 | 85 (15%) | 50 (13%) |  |
| 15-24 | 190 (34%) | 128 (34%) |  |
| 25-39 | 177 (31%) | 124 (33%) |  |
| 40-59 | 68 (12%) | 47 (13%) |  |
| >59 | 28 (5.0%) | 16 (4.3%) |  |
| **Occupation** |  |  |  |
| Minor | 24 (4.2%) | 15 (4.0%) | 0.68 |
| Student | 130 (23%) | 82 (22%) |  |
| Agricultural | 243 (43%) | 175 (46%) |  |
| Other | 55 (9.6%) | 15 (4.0%) |  |
| Unemployed | 118 (21%) | 66 (17%) |  |
| **Nationality** |  |  |  |
| Eswatini | 478 (84%) | 303 (80%) | 0.18 |
| Mozambique | 94 (16%) | 76 (20%) |  |
| **Travel** |  |  |  |
| No Travel | 385 (67%) | 245 (65%) | 0.82 |
| Domestic | 46 (8.1%) | 28 (7.4%) |  |
| Mozambique | 127 (22%) | 95 (25%) |  |
| Other Country | 11 (1.9%) | 9 (2.4%) |  |
| Multiple | 2 (0.4%) | 2 (0.5%) |  |
| **Detection method** |  |  | 0.82 |
| Passive | 538 (94%) | 353 (93%) |  |
| Active | 2 (0.3%) | 2 (0.5%) |  |
| Reactive | 32 (5.6%) | 24 (6.3%) |  |
| **Region** |  |  | 0.98 |
| Hhohho | 292 (51%) | 196 (52%) |  |
| Lubombo | 209 (37%) | 134 (35%) |  |
| Manzini | 62 (11%) | 42 (11%) |  |
| Shiselweni | 9 (1.6%) | 7 (1.8%) |  |

*Samples with a valid genotype in all 59 assessed *kelch13* SNPs

**chi-squared test

**Table S2. Comparison of characteristics and origin of recruited participants who traveled to Mozambique to those who did not travel or traveled to other countries.** This table only includes samples with valid genotypes in all 59 assessed *kelch13* SNPs

|  | **No travel to Mozambique** | **Travel to Mozambique** | ***P*-value*** |
| --- | --- | --- | --- |
| ***N*** | 293 | 98 |  |
| **Sex** |  |  | 0.005 |
| Female | 75 (26%) | 11 (11%) |  |
| Male | 218 (74%) | 87 (89%) |  |
| **Age** |  |  | 0.06 |
| <5 | 10 (3.5%) | 2 (2.0%) |  |
| 5-14 | 45 (15%) | 5 (5.1%) |  |
| 15-24 | 92 (32%) | 42 (43%) |  |
| 25-39 | 97 (34%) | 29 (30%) |  |
| 40-59 | 33 (11%) | 15 (15%) |  |
| >59 | 11 (3.8%) | 5 (5.1%) |  |
| Unknown | 3 | 0 |  |
| **Occupation** |  |  | <0.001 |
| Minor | 13 (4.5%) | 3 (3.1%) |  |
| Student | 81 (28%) | 3 (3.1%) |  |
| Agricultural | 113 (39%) | 66 (67%) |  |
| Other | 19 (6.6%) | 22 (22%) |  |
| Unemployed | 64 (22%) | 4 (4.1%) |  |
| Unknown | 1 | 0 |  |
| **Detection method** |  |  | 0.71 |
| Passive | 271 (93%) | 92 (94%) |  |
| Active | 2 (0.7%) | 0 (0%) |  |
| Reactive | 18 (6.2%) | 6 (6.1%) |  |
| **Region** |  |  | 0.002 |
| Hhohho | 149 (51%) | 49 (50%) |  |
| Lubombo | 113 (39%) | 27 (28%) |  |
| Manzini | 23 (7.9%) | 21 (21%) |  |
| Shiselweni | 6 (2.1%) | 1 (1.0%) |  |

*chi-squared test

**Table S3. Logistic regression of infection polyclonality and case characteristics.**

|  | **Univariate logistic regression** | | | **Multivariate logistic regression** | | |
| --- | --- | --- | --- | --- | --- | --- |
|  | ***n*/*N*** | **OR (95% CI)** | ***P*-value** | ***n*/*N* (%)** | **OR (95% CI)** | ***P*-value** |
| **Age** |  |  |  |  |  |  |
| <5 | 7/14 (50%) | Ref. |  | 7/14  (50%) | Ref. |  |
| 5-14 | 37/59  (62.7%) | 1.68  (0.51-5.55) | 0.4 | 37/59  (62.7%) | 1.78  (0.24-17.1) | 0.6 |
| 15-24 | 98/154  (63.6%) | 1.75  (0.57-5.36) | 0.3 | 98/153  (62.7%) | 3.13 (0.35-35.0) | 0.3 |
| 25-39 | 81/149  (54.4%) | 1.19  (0.39-3.64) | 0.8 | 77/144  (53.5%) | 2.53 (0.27-29.7) | 0.4 |
| 40-59 | 29/52  (55.8%) | 1.26  (0.38-4.19) | 0.7 | 28/51  (54.9%) | 2.93 (0.29-36.0) | 0.4 |
| 60+ | 10/20  (50%) | 1.00  (0.25-3.98) | >0.9 | 10/20  (50.0%) | 2.99 (0.26-40.8) | 0.4 |
| **Sex** |  |  |  |  |  |  |
| Female | 66/106  (62.3%) | Ref. |  | 61/101  (60.4%) | Ref. |  |
| Male | 201/350  (57.4%) | 0.82  (0.52-1.27) | 0.4 | 196/340  (57.6%) | 0.96  (0.56-1.66) | 0.9 |
| **Occupation** |  |  |  |  |  |  |
| Minor | 11/20  (55%) | Ref. |  | 11/20  (55%) | Ref. |  |
| Student | 61/95  (64.2%) | 1.47  (0.54-3.90) | 0.4 | 60/92  (65.2%) | 0.81  (0.10-4.87) | 0.8 |
| Agricultural | 121/209  (57.9%) | 1.13  (0.44-2.83) | 0.8 | 119/206  (57.8%) | 0.35 (0.04-2.45) | 0.3 |
| Other | 26/45  (57.8%) | 1.12  (0.38-3.24) | 0.9 | 25/43  (78.1%) | 0.30 (0.03-2.34) | 0.3 |
| Unemployed | 45/83  (54.2%) | 0.97  (0.36-2.59) | >0.9 | 42/80  (52.5%) | 0.31  (0.03-2.29) | 0.3 |
| **Detection method** |  |  |  |  |  |  |
| Passive | 247/427  (57.8%) | Ref. |  | 225/384  (58.6%) | Ref. |  |
| Proactive and reactive | 16/25  (64.0%) | 1.30  (0.57-3.12) | 0.5 | 18/27  (66.7%) | 1.01 (0.42-2.55) | 0.8 |
| **Travel History** |  |  |  |  |  |  |
| No Travel | 162/300  (54.0%) | Ref. |  | 154/289  (53.3%) | Ref. |  |
| Domestic | 22/36  (61.1%) | 1.34  (0.67-2.77) | 0.4 | 22/36  (61.1%) | 1.19 (0.55-2.67) | 0.7 |
| Mozambique | 78/108  (72.2%) | 2.21  (1.38-3.61) | 0.001 | 78/107  (72.9%) | 2.51  (1.45-4.45) | 0.001 |
| Other | 3/9  (33.3%) | 0.43  (0.09-1.65) | 0.2 | 3/9  (33.3%) | 0.48 (0.09-2.20) | 0.4 |
| **Parasitemia** |  |  |  |  |  |  |
| log10(parasite density) | N=456 | 0.64 (0.54-0.75) | <0.001 | N=441 | 0.72 (0.59-0.86) | <0.001 |
| **Depth of coverage** |  |  |  |  |  |  |
| log10(Reads) | N=456 | 0.42  (0.29-0.60) | <0.001 | N=441 | 0.58  (0.39-0.87) | 0.009 |

**Table S4. Proportion of samples carrying a *dhps* or *dhfr* haplotype.**

n: number of samples with the haplotype. N: total number of samples with a valid haplotype. CI: confidence interval.

| *n*/*N*  %  [95% CI] | Overall | Hhohho | Lubombo | Manzini | Shiselweni |
| --- | --- | --- | --- | --- | --- |
| ***dhps*** |  |  |  |  |  |
| ISAKAA^a^ | 10/429 2.3% [1.1-4.2%] | 5/227  2.2%  [0.7-5.1%] | 4/149  2.7%  [0.7-6.7%] | 0/47  0% | 1/6  16.7%  [0.4-64.1%] |
| ISGKAA | 8/429 1.9% [0.8-3.6%] | 2/227  0.9%  [0.1-3.1%] | 3/149  2.0%  [0.4-5.8%] | 3/47  6.4%  [1.3-17.5%] | 0/6  0% |
| ISAEAA | 3/429 0.7% [0.1-2%] | 0/227  0% | 2/149  1.3%  [0.2-4.8%] | 1/47  2.1%  [0.1-11.3%] | 0/6  0% |
| ISGEAA^b^ | 405/429 94.4% [91.8-96.4%] | 219/227  96.5%  [93.2-98.5%] | 140/149  94.0%  [88.8-97.2%] | 42/47  89.4%  [76.9-96.5%] | 4/6  66.7%  [22.3-95.7%] |
| ISGEGA^c^ | 29/429 6.8% [4.6-9.6%] | 10/227  4.4%  [2.1-8.0%] | 13/149  8.7%  [4.7-14.5%] | 6/47  12.8%  [4.8-25.7%] | 0/6  0% |
| VAGKGS | 3/429 0.7% [0.1-2%] | 1/227  0.4%  [0.0-2.4%] | 0/141  0% | 1/47  2.1%  [0.1%] | 1/6  16.7%  [0.4-64.1%] |
| Undetermined^d^ | 22 | 6 | 13 | 2 | 1 |
| ***dhfr*** |  |  |  |  |  |
| NCSI | 1/453 0.2% [0-1.2%] | 1/235 0.4% [0-2.3%] | 0/162 0% | 0/48 0% | 0/7 0% |
| NRNI | 20/453 4.4% [2.7-6.7%] | 5/235 2.1% [0.7-4.9%] | 13/162 8.0% [4.3-13.3%] | 2/49 4.2% [0.5-14.0%] | 0/7 0% |
| IRNI^e^ | 448/453 98.9% [97.4-99.6%] | 232/235 98.7% [96.3-99.7%] | 160/162 98.8% [95.6-99.9%] | 49/49 100% [92.7-100%] | 7/7 100% [59-100%] |
| Undetermined^d^ | 1 | 1 | 0 | 0 | 0 |

**Table S5. Logistic regression of *dhps*/*dhfr* quintuple mutant carriage and case characteristics.**

|  | **Univariate logistic regression** | | | **Multivariate logistic regression** | | |
| --- | --- | --- | --- | --- | --- | --- |
|  | ***n*/*N* (%)** | **OR (95% CI)** | ***P*-value** | ***n*/*N* (%)** | **OR (95% CI)** | ***P*-value** |
| **Age** |  |  |  |  |  |  |
| <15 | 64/72 (88.9%) | Ref. |  | 64/72 (88.9%) | Ref. |  |
| 15-24 | 142/155 (91.6%) | 1.37 (0.52-3.40) | 0.5 | 141/154 (91.6%) | 1.44 (0.40-5.38) | 0.6 |
| 25-39 | 122/143 (85.3%) | 0.73 (0.29-1.67) | 0.5 | 117/138 (84.8%) | 0.52 (0.11-2.35) | 0.4 |
| 40+ | 64/73 (87.7%) | 0.89 (0.32-2.47) | 0.8 | 63/72 (87.5%) | 1.29 (0.25-6.89) | 0.8 |
| **Sex** |  |  |  |  |  |  |
| Female | 90/106 (84.9%) | Ref. |  | 85/101 (84.2%) | Ref. |  |
| Male | 310/345 (89.9%) | 1.57 (0.81-2.93) | 0.2 | 300/335 (89.6%) | 1.1 (0.50-2.35) | 0.8 |
| **Occupation** |  |  |  |  |  |  |
| Minor | 19/20 (95%) | Ref. |  | 19/20 (95%) | Ref. |  |
| Student | 81/93 (87.1%) | 0.36 (0.02-1.98) | 0.3 | 78/90 (86.7%) | 0.33 (0.02-2.12) | 0.3 |
| Agricultural | 192/207 (92.8%) | 0.67 (0.04-3.62) | 0.7 | 189/204 (92.6%) | 1.03 (0.04-10.1) | >0.9 |
| Other | 39/45 (86.7%) | 0.34 (0.02-2.20) | 0.3 | 37/43 (86%) | 0.58 (0.02-6.89) | 0.7 |
| Unemployed | 65/82 (79.3%) | 0.2 (0.01-1.08) | 0.13 | 62/79 (78.5%) | 0.18 (0.01-1.75) | 0.2 |
| **Detection method** |  |  |  |  |  |  |
| Passive | 374/422 (88.6%) | Ref. |  | 363/411 (88.3%) | Ref. |  |
| Proactive and reactive | 22/25 (88%) | 0.94 (0.31-4.08) | >0.9 | 22/25 (88%) | 1.2 (0.36-5.78) | 0.8 |
| **Travel History** |  |  |  |  |  |  |
| No Travel | 264/294 (89.8%) | Ref. |  | 253/283 (89.4%) | Ref. |  |
| Domestic | 34/37 (91.9%) | 1.29 (0.43-5.57) | 0.7 | 34/37 (91.9%) | 1.65 (0.50-7.55) | 0.5 |
| Mozambique | 96/108 (88.9%) | 0.91 (0.46-1.91) | 0.8 | 95/107 (88.8%) | 0.51 (0.21-1.23) | 0.13 |
| Other | 3/9 (33.3%) | 0.06 (0.01-0.23) | <0.001 | 3/9 (33.3%) | 0.05 (0.01-0.23) | <0.001 |
| **Parasitemia** |  |  |  |  |  |  |
| log10(Reads) | N=449 | 1.03 (0.82-1.28) | 0.8 | N = 434 | 1.07 (0.83-1.36) | 0.6 |

**Table S6. Logistic regression of *dhps*/*dhfr* sextuple mutant carriage and case characteristics.**

|  | **Univariate logistic regression** | | | **Multivariate logistic regression** | | |
| --- | --- | --- | --- | --- | --- | --- |
|  | ***n*/*N* (%)** | **OR (95% CI)** | ***P*-value** | ***n*/*N* (%)** | **OR (95% CI)** | ***P*-value** |
| **Age** |  |  |  |  |  |  |
| <15 | 4/72 (5.6%) | Ref. |  | 4/71 (5.6%) | Ref. |  |
| 15-24 | 11/155 (7.1%) | 1.3 (0.43-4.82) | 0.7 | 11/151 (7.3%) | 1.64 (0.52-6.30) | 0.4 |
| 25-39 | 11/143 (7.7%) | 1.42 (0.46-5.26) | 0.6 | 11/135 (8.1%) | 1.7 (0.54-6.53) | 0.4 |
| 40+ | 2/73 (2.7%) | 0.48 (0.06-2.54) | 0.4 | 2/70 (2.9%) | 0.63 (0.08-3.42) | 0.6 |
| **Sex** |  |  |  |  |  |  |
| Female | 9/106 (8.5%) | Ref. |  | 8/99 (8.1%) | Ref. |  |
| Male | 20/345 (5.8%) | 0.66 (0.30-1.58) | 0.3 | 20/328 (6.1%) | 0.79 (0.33-2.05) | 0.6 |
| **Travel History** |  |  |  |  |  |  |
| No Travel | 24/294 (8.2%) | Ref. |  | 23/283 (8.1%) | Ref. |  |
| Domestic | 2/37 (5.4%) | 0.64 (0.10-2.30) | 0.6 | 2/37 (5.4%) | 0.59 (0.09-2.19) | 0.5 |
| Mozambique | 3/108 (2.8%) | 0.32 (0.08-0.94) | 0.069 | 3/107 (2.8%) | 0.29 (0.07-0.89) | 0.053 |
| **Parasitemia** |  |  |  |  |  |  |
| log10(Reads) | N=449 | 0.81 (0.62-1.07) | 0.13 | N = 429 | 0.78 (0.59-1.06) | 0.1 |

**Table S7. Logistic regression of *mdr1* F184Y and case characteristics.**

|  | **Univariate logistic regression** | | | **Multivariate logistic regression** | | |
| --- | --- | --- | --- | --- | --- | --- |
|  | ***n*/*N* (%)** | **OR (95% CI)** | ***P*-value** | ***n*/*N* (%)** | **OR (95% CI)** | ***P*-value** |
| **Age** |  |  |  |  |  |  |
| <5 | 10/13 (76.9%) | Ref. |  | 10/13 (76.9%) | Ref. |  |
| 5-14 | 35/58 (60.3%) | 0.46 (0.09-1.68) | 0.3 | 35/58 (60.3%) | 0.14 (0.01-1.09) | 0.073 |
| 15-24 | 80/149 (53.7%) | 0.35 (0.08-1.19) | 0.12 | 80/148 (54.1%) | 0.1 (0.01-0.94) | 0.052 |
| 25-39 | 83/140 (59.3%) | 0.44 (0.09-1.50) | 0.2 | 78/135 (57.8%) | 0.14 (0.01-1.34) | 0.1 |
| 40-59 | 30/53 (56.6%) | 0.39 (0.08-1.45) | 0.2 | 29/52 (55.8%) | 0.11 (0.01-1.15) | 0.075 |
| 60+ | 13/20 (65%) | 0.56 (0.10-2.58) | 0.5 | 13/20 (65%) | 0.18 (0.01-2.15) | 0.2 |
| **Sex** |  |  |  |  |  |  |
| Female | 59/100 (59%) | Ref. |  | 56/96 (58.3%) | Ref. |  |
| Male | 195/340 (57.4%) | 0.93 (0.59-1.47) | 0.8 | 189/330 (57.3%) | 0.99 (0.58-1.67) | >0.9 |
| **Occupation** |  |  |  |  |  |  |
| Minor | 12/19 (63.2%) | Ref. |  | 12/19 (63.2%) | Ref. |  |
| Student | 55/93 (59.1%) | 0.84 (0.29-2.30) | 0.7 | 55/91 (60.4%) | 4.08 (0.71-32.0) | 0.13 |
| Agricultural | 114/202 (56.4%) | 0.76 (0.27-1.96) | 0.6 | 111/199 (55.8%) | 2.84 (0.43-24.7) | 0.3 |
| Other | 27/42 (64.3%) | 1.05 (0.33-3.21) | >0.9 | 25/40 (62.5%) | 3.54 (0.48-33.6) | 0.2 |
| Unemployed | 44/80 (55%) | 0.71 (0.24-1.96) | 0.5 | 42/77 (54.5%) | 2.96 (0.43-26.5) | 0.3 |
| **Detection method** |  |  |  |  |  |  |
| Passive | 234/411 (56.9%) | Ref. |  | 228/401 (56.9%) | Ref. |  |
| Proactive and reactive | 17/25 (68%) | 1.61 (0.70-4.02) | 0.3 | 17/25 (68%) | 1.75 (0.74-4.50) | 0.2 |
| **Travel History** |  |  |  |  |  |  |
| No Travel | 158/287 (55.1%) | Ref. |  | 152/277 (54.9%) | Ref. |  |
| Domestic | 22/34 (64.7%) | 1.5 (0.72-3.23) | 0.3 | 22/34 (64.7%) | 1.55 (0.72-3.45) | 0.3 |
| Mozambique | 68/107 (63.6%) | 1.42 (0.90-2.26) | 0.13 | 68/106 (64.2%) | 1.68 (1.00-2.87) | 0.054 |
| Other | 3/9 (33.3%) | 0.41 (0.08-1.58) | 0.2 | 3/9 (33.3%) | 0.33 (0.07-1.35) | 0.14 |
| **Parasitemia** |  |  |  |  |  |  |
| log10(parasite density) | N=430 | 1 (0.87-1.16) | >0.9 | N=426 | 1.06 (0.91-1.24) | 0.4 |

**Table S8. Logistic regression of *mdr2* I492V and case characteristics.**

|  | **Univariate logistic regression** | | | **Multivariate logistic regression** | | |
| --- | --- | --- | --- | --- | --- | --- |
|  | ***n*/*N* (%)** | **OR (95% CI)** | ***P*-value** | ***n*/*N* (%)** | **OR (95% CI)** | ***P*-value** |
| **Age** |  |  |  |  |  |  |
| <5 | 2/11 (18.2%) | Ref. |  | 2/11 (18.2%) | Ref. |  |
| 5-14 | 12/51 (23.5%) | 1.38 (0.30-9.90) | 0.7 | 12/51 (23.5%) | 1.4 (0.05-21.3) | 0.8 |
| 15-24 | 27/138 (19.6%) | 1.09 (0.26-7.45) | >0.9 | 27/138 (19.6%) | 1.08 (0.04-19.5) | >0.9 |
| 25-39 | 39/127 (30.7%) | 1.99 (0.49-13.5) | 0.4 | 39/122 (32%) | 2.16 (0.07-41.2) | 0.6 |
| 40-59 | 15/45 (33.3%) | 2.25 (0.50-16.0) | 0.3 | 15/44 (34.1%) | 2.64 (0.08-52.9) | 0.5 |
| 60+ | 5/15 (33.3%) | 2.25 (0.38-18.6) | 0.4 | 5/15 (33.3%) | 2.79 (0.08-65.8) | 0.5 |
| **Sex** |  |  |  |  |  |  |
| Female | 18/83 (21.7%) | Ref. |  | 18/81 (22.2%) | Ref. |  |
| Male | 82/309 (26.5%) | 1.3 (0.74-2.38) | 0.4 | 82/300 (27.3%) | 1.21 (0.63-2.39) | 0.6 |
| **Occupation** |  |  |  |  |  |  |
| Minor | 3/15 (20%) | Ref. |  | 3/15 (20%) | Ref. |  |
| Student | 18/85 (21.2%) | 1.07 (0.30-5.08) | >0.9 | 18/83 (21.7%) | 0.91 (0.10-19.6) | >0.9 |
| Agricultural | 53/184 (28.8%) | 1.62 (0.49-7.31) | 0.5 | 53/181 (29.3%) | 0.88 (0.08-20.6) | >0.9 |
| Other | 11/39 (28.2%) | 1.57 (0.40-7.86) | 0.5 | 11/37 (29.7%) | 0.73 (0.06-18.5) | 0.8 |
| Unemployed | 15/66 (22.7%) | 1.18 (0.32-5.65) | 0.8 | 15/65 (23.1%) | 0.67 (0.06-16.6) | 0.8 |
| **Detection method** |  |  |  |  |  |  |
| Passive | 93/365 (25.5%) | Ref. |  | 93/358 (26%) | Ref. |  |
| Proactive and reactive | 7/23 (30.4%) | 1.28 (0.48-3.10) | 0.6 | 7/23 (30.4%) | 1.21 (0.44-3.02) | 0.7 |
| **Travel History** |  |  |  |  |  |  |
| No Travel | 58/253 (22.9%) | Ref. |  | 58/245 (23.7%) | Ref. |  |
| Domestic | 7/29 (24.1%) | 1.07 (0.41-2.52) | 0.9 | 7/29 (24.1%) | 1.05 (0.38-2.63) | >0.9 |
| Mozambique | 31/99 (31.3%) | 1.53 (0.91-2.56) | 0.1 | 31/99 (31.3%) | 1.37 (0.75-2.49) | 0.3 |
| Other | 4/8 (50%) | 3.36 (0.77-14.6) | 0.093 | 4/8 (50%) | 3.55 (0.78-16.4) | 0.092 |
| **Parasitemia** |  |  |  |  |  |  |
| log10(parasite density) | N=392 | 0.9 (0.76-1.07) | 0.2 | N=381 | 0.87 (0.72-1.05) | 0.13 |
